# Interplay of Generation Time and Spatial Structure in Epidemic Dynamics and the Reliability of Reproduction Ratio Estimates

**DOI:** 10.1101/2025.09.02.25334936

**Authors:** Claudio Ascione, Eugenio Valdano

## Abstract

The reproduction ratio *R* is a central metric for monitoring infectious disease epidemics and guiding public health interventions. It is typically inferred from population-level surveillance data, but such estimates can be biased by the spatial structure of the underlying population and by complexities in disease natural history. Here, we develop a theoretical framework to study how the distribution of the generation time (the time from primary to secondary infection) interacts with spatial network structure of the host population, to shape epidemic dynamics and the accuracy of *R* estimates. We show that the mean and dispersion of the generation time determine the contribution of subdominant epidemic modes, controlling the rate at which the system converges to its spatial equilibrium. Overdispersed generation times slow convergence near the epidemic threshold, whereas underdispersed distributions, common for respiratory pathogens, can markedly delay convergence at moderate and high transmissibility, producing long-lived biases in *R*. We evaluate the performance of an existing correction to incidence data that removes bias from spatial structure under simpified dynamical conditions. We demonstrate that it remains valid for arbitrary generation time distributions only if both mean and dispersion are accurately specified. Otherwise, substantial residual errors persist especially when *R* is above threshold. We extend the framework to short-lived perturbations, both exogenous (e.g., extreme weather, mass gatherings, mobility restrictions) and endogenous (e.g., behavioral changes), and show that realistic dispersions can amplify their impact on spatial distributions and prolong bias on *R* far beyond the perturbation. We illustrate these mechanisms through a case study of a respiratory pathogen in Spain, using colocation-based mobility data, and identify the conditions under which surveillance-derived *R* is reliable and when precise measurements of the generation time distribution are critical.

## 1 Introduction

The reproduction ratio *R* is a widely used indicator for monitoring the evolution of an infectious disease epidemic [1, 2]. It quantifies the average number of secondary infections generated by one infected individual over the course of the infection. Estimates of the reproduction ratio underpin public health decisions and interventions, as they allow authorities to assess pathogen circulation and compare alternative scenarios [3–5].

The reproduction ratio is inferred, across a wide range of diseases and epidemics [5–9], from population-level data coming from epidemiological surveillance, such as incident cases, healthcare access (e.g., hospitalizations), or severe outcomes [10–12]. Such estimates, however, are sensitive both to the underlying epidemic dynamics, and to the accuracy of the available data and of the measurements that are made on those data. Specifically, the reliability of the estimated value of *R* is known to depend on the underlying parametrization of the distribution of the generation time [10, 13, 14]. The generation time is the time between the infection of an index case and the time at which that case infects someone else. The generation time, and its empirical distribution, is, too, typically inferred from epidemiological data [15–17]. Different diseases exhibit different generation time distributions, and even under similar epidemic conditions, these differences can lead to substantially different reproduction ratio estimates [18]. At the same time, in a recent study we showed that the estimates of *R* obtained through statistical inference on surveillance data are biased if one does not properly account for the spatial structure of the host population on which the disease spread [19]. This is due to the complex network structures describing contacts and emerging from host demographic and behavioral and mobility patterns within and across spatial communities [20–25].

Available theoretical methods for estimating *R* from surveillance, however, either control for the heterogeneity in the generation time in spatially homogeneous populations [10], or address the complexity of the spatial structure but assume a fixed generation time [19]. Here, we study the interaction between these two aspects and analytically determine when they can be treated separately, and when instead their complex interplay has a nontrivial effect on epidemic dynamics and consequently the estimate of the reproduction ratio. Finally, we use the developed theory to study the effects of temporally localized perturbations on spatial epidemics dynamics and their effect on the estimate of epidemic trends. This applies to commonly observed phenomena such as extreme weather and natural disasters [26–29], public health interventions such as mobility restrictions [30–32], seasonal patterns such as holidays [33, 34].

## 2 Theoretical formalism

We consider a population made of *N* distinct spatial communities. Let *I*_*i*_(*t*) be the number of incident (new) infections occurring among residents of community *i* during an infinitesimal time interval centered in *t*. Then, let *ζ* be the probability density function of the generation time: *ζ*(*t*) *≥* 0 and 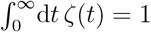. **R** is the *N × N* reproduction operator as defined in Ref. [19]. This operator is similar to the well-known next-generation matrix [24, 35, 36] and encodes the transmission potential of the disease within and between communities. Specifically, *R*_*ij*_ is the expected number of secondary infections that a resident of *j* generates over the course of their infectious period among residents of *i*. The reproduction ratio of the system, called *reference* reproduction ratio henceforth following the nomenclature of Ref. [19], is then the largest eigenvalue of *R*_ref_ = *ρ*[**R**] [37, 38], where *ρ*[*·*] is the spectral radius of the operator. This value is real and positive because **R** is nonnegative and can be assumed irreducible [19]. Then, the evolution of the number of infections obeys the following Volterra integral equation:

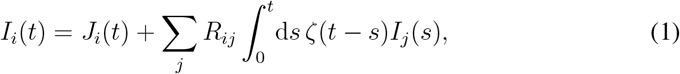

where *J*_*i*_(*t*) is a source term accounting for importation and exportation of infections from outside of the system. This modeling approach has been widely used [1, 39–41] and offer the advantage of being able to consider completely general generation time distributions. It discounts, however, stochastic fluctuations and assumes that **R** remains approximately constant during the period in which we study the epidemic: this is a reasonable assumption if we are in the early phase of an epidemic when no acquired immunity or major public health interventions have been put in place. It is also reasonable whenever the time scale of observation is shorter than typical time scales such as seasonality, vital dynamics and immunity buildup and waning [19, 42].

## 3 Evolution of the dynamics

Eq. (1) admits the following as a general solution [43]:

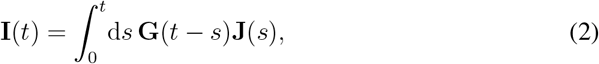

which we wrote in vector notation where **I** is simply the vector with *I*_*i*_ as components. The operator **G**(*t*) is the system Green’s function which solves the following equation:

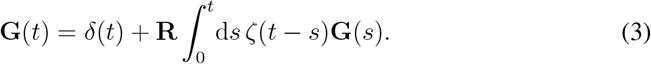

Equation (3) admits a formal solution in Laplace space [44]:

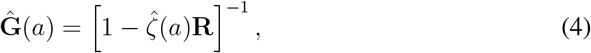

where ^ denotes the Laplace transform and *a* the variable in the Laplace space. It can be written in the basis of eigenvectors of **R**:

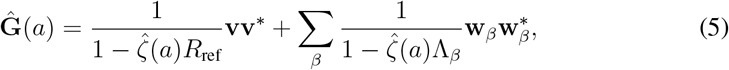

where **v** is the (right) Perron eigenvector associated to *R*_ref_, **v**^*\**^ the corresponding covector in the conjugate base (left eigenvector), Λ_*β*_ are the other *N −* 1 eigenvalues of **R** (repeated according to their multiplicities) and **w**_*β*_, 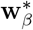 the corresponding eigenvectors and their conjugates. Vectors are normalized so that ∑_*i*_ *v*_*i*_ = 1, **v**^*\**^**v** = 1, 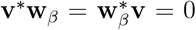 and 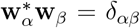. We can determine the evolution in time of **G**(*t*) from the singularities of **Ĝ** (*a*) in Eq. (5). Asymptotically at large time it will evolve as 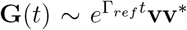 where Γ_*ref*_ is the solution of 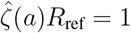 with largest real part (it will be itself real and positive) – see Appendix A for details on the large-time evolution. This fact is a generalization of the link between the basic reproduction ratio and the epidemic growth rate which is well-known in the context of populations without spatial structure [10]. At shorter times other modes will contribute, too. Some come from other possible solutions of 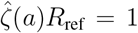, other from the subdominant modes of the spectral decomposition of **R**: they are in the form 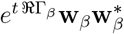 where Γ_*β*_ is a solution of 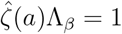.

These considerations confirm, on the one hand, that in the large-time limit the spatial distribution of infections will reach an equilibrium distribution corresponding to **v**, which was previously proven by assuming a constant generation time [19]. On the other hand, they show that the specific generation time distribution may strongly affect both the pathway and the rate at which the epidemic converges to that equilibrium distribution, by altering the relative rates of decay of the subdominant modes with respect to the leading one – which in turn determine our ability to estimate *R*_ref_ from surveillance.

## 4 Estimating the reproduction ratio from surveillance data

As a first step, we study how the generation time distribution combined with the spatial network structure affect the estimate of the reproduction ratio. We define *I*_*tot*_(*t*) =Σ_*i*_ *I*_*i*_(*t*) as the total incidence of infections and we also define the corrected incidence 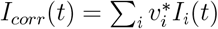, as it was introduced in Ref. [19], to remove the bias on the estimate of the reproduction ratio induced by the spatial structure when the generation time was assumed constant. We also introduce *ζ*_*estim*_ as the estimated generation time distribution, which is the one specified when estimating *R*_ref_. The generation time distribution may be correctly specified (*ζ*_*estim*_ = *ζ*) or not, if the estimated mean and/or dispersion do not match the true values. It is typically the parametrization of the generation time which is available either from similar epidemic contexts or as a best estimate on available data. We call *R*_estim_(*t*) and *R*_corr_(*t*) the reproduction ratios estimated from *I*_*tot*_ and *I*_*corr*_, respectively. The former (*R*_estim_(*t*)) is the surveillance-based reproduction ratio as customarily estimated by public health autorities without correcting from the spatial structure. Within our framework, they should instantly solve the following effective equations:

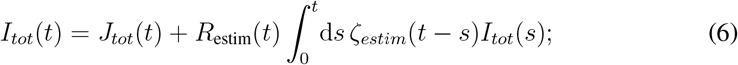

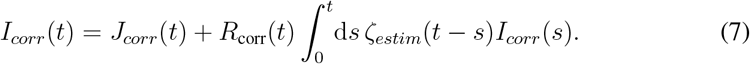

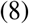

### 4.1 When the generation time is correctly specified

First, we will consider the case when the generation time, and its distribution, are accurately estimated and specified: *ζ*_*estim*_ = *ζ*. Applying **v**^*\**^ on both sides of Eq. (1) one gets

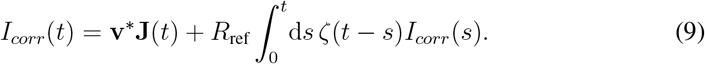

Comparing it with Eq. (7), this means that, if importation and exportation of cases are negligible, then *R*_corr_(*t*) = *R*_ref_ at all times: the corrected incidence introduced in Ref. [19] still correctly removes the bias induced by the spatial network provided that the generation time is known and movement of cases in and out of the system is negligible. If the latter condition does not hold, then importation and exportation will bias the estimate of the reproduction ratio with a factor proportional to the projection of the source term **J** onto the equilibrium spatial distribution:

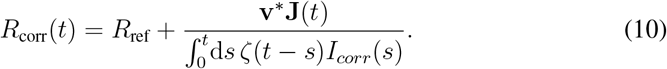

Given that both the equilibrium distribution and the hubs of case importation and exportation tend to concentrate in densely populated communities, this source of bias may be relevant especially at the start of the epidemic. At that time, the incidence of internally generated infections is low and possibly comparable to introductions, and typically hard to control for as the form of **J**(*t*) is poorly known [45–47].

To study the behavior of *R*_estim_, we adopt the common approach of modeling the generation time distribution with a gamma distribution, parametrized in terms of its mean value *τ*, representing the average generation time, and the shape parameter *k* tuning the dispersion around the mean. *k* = 1 is an exponential distribution, with variance equal to the mean squared. An exponentially-distributed generation time is common to the simplest compartmental models such as the Susceptible-Infectious-Susceptible (SIS) or the Susceptible-Infectious-Recovered (SIR). Values of *k >* 1 give distributions that are underdispersed with respect to the exponential distribution (variance smaller than *τ* ^2^) while *k <* 1 are overdispersed (variance larger than *τ* ^2^) and have broader tails than the exponential. More on this parametrization is available in Appendix B. Estimates from observational data often lead to underdispersed distribution (*k >* 1): Figure 1(A,C) shows the generation time distribution of COVID-19 (original strain in 2020) and pandemic H1N1 influenza from 2009, as estimated in Ref. [48, 49], compared to alternative dispersion (*k*) values. Cases of overdispersed generation times, albeit rarer, have also been reported [50].

**Fig.1.**
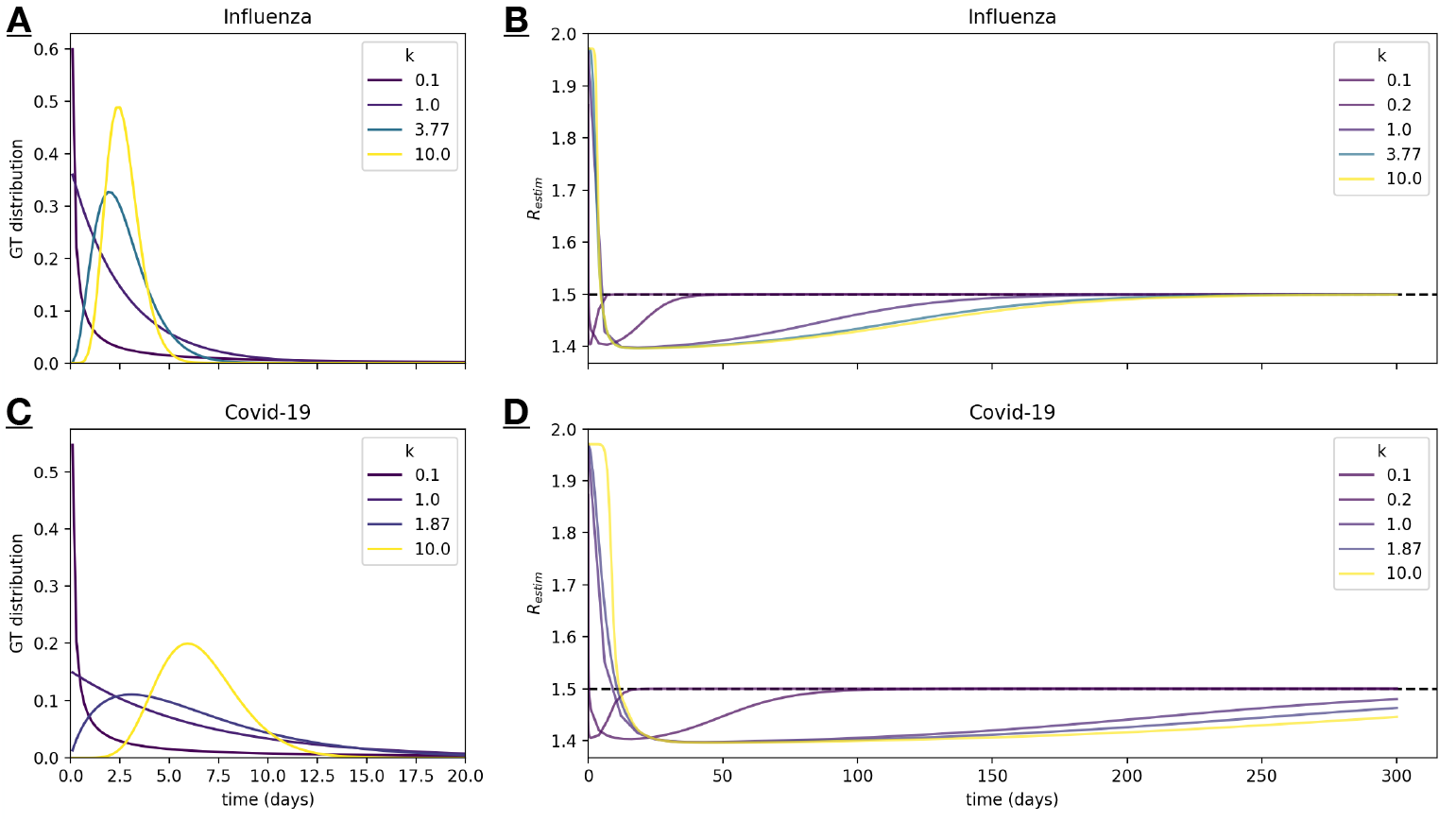
In panels **A** and **C**, a gamma distribution is shown with the mean set to the estimated values for COVID-19 (original strain) [49] and the H1N1 influenza pandemic [48], respectively, while varying the shape parameter *k*. Panels **B** and **D** show the temporal evolution of *R*_estim_, computed assuming a generation time distribution modeled as a gamma distribution with the same range of *k* values used respectively in panels **A** and **C**. *R*_ref_ is represented as a dotted line.

Using the Laplace transform (see Appendix B) we can establish the time evolution of **G**(*t*) through the exponential growth rates coming from each eigenmode of the spatial network **v, w**_*β*_:

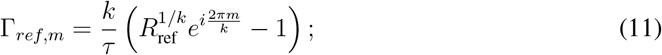

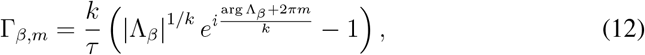

where we have specified arg Λ_*β*_ to account for the fact that some eigenvalues of **R** may be negative or nonreal [19]. Appendix B proves that, for each mode, *m* runs over a finite set of indices, being *{*0, 1, *· · ·, k −* 1*}* if *k ∈*ℕ or one subset of it if not. It also proves that the full dynamical evolution can be written as

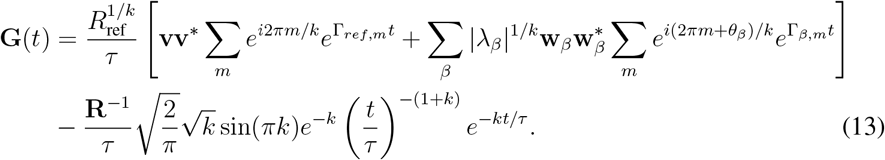

If *k ∈*ℕ, Eq. (13) is exact and is the generalization to a spatially structured population of the series representation available in Ref. [41]. If *k∉*ℕ, the last term is nonzero and is an additional contribution to each eigenmode, coming from the branch point of the generation-time distribution. This contribution always decays in time (even above the epidemic threshold), and its weight is suppressed for under-dispersed generation times (*k* ≳ 2). Nevertheless, it can become significant in the presence of very weak eigenmodes (small Λ_*β*_) or for highly over-dispersed generation times (*k ≈* 1*/*2). Further details on the computation of this and the other components of Eq. (13) are provided in Appendix B.

The term 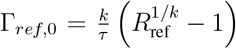 dominates the asymptotic (large time) dynamics of the system, and represents the well-known relationship between the reproduction ratio and the system asymptotic growth rate for a gamma-distributed generation time [10, 41]. The limit *k* = 1 gives the familiar value of Γ_*ref*,0_ = (*R*_ref_ *−* 1)*/τ* of the SIR and SIS models, and the limit *k → ∞* gives Γ_*ref*,0_ = log *R*_ref_*/τ*, which corresponds to the case of fixed generation time of Ref. [19]. The other modes Γ_*ref,m*_ and Γ_*β,m*_ are subdominant and decay relative to Γ_*ref*,0_: the speed at which they decay determines the accuracy of *R*_estim_ [19]. This is because Eq. (13) describes the interaction between the spatial network linking the different communities and the generation time distribution in determining the epidemic dynamics and, as such, the accuracy in estimating the reproduction ratio. To study this dynamics, we can study the weight of each eigenmode and its speed of decay from Eq. (13). This shows that each subdominant eigenmode (barring the additional, *k*-suppressed one) of the spatial network contributes proportionally to |*λ*_*β*_|^1*/k*^ (where *λ*_*β*_ = Λ_*β*_*/R*_ref_), which is a growing function of *k* given that |*λ*_*β*_| *<* 1. Two consequences follow. First, decreasing the dispersion of the generation time around its mean (i.e., increasing *k*) increases the weights of the subdominant spatial eigenmodes. Second, the relative difference between the weights of two distincts eigenmodes will also decrease as *k* grows, meaning that more eigenmodes may give a perceptible contribution to the epidemic evolution. Even more crucially, the generation time distribution impacts the rate at which the subdominant eigenmodes decay relative to the dominant one. Using Eq. (12) we can define such relative rate as follows:

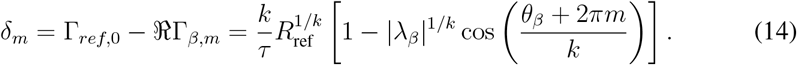

Without loss of generality we will consider now a generic

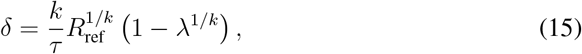

thus assuming *λ* real and positive. If not, it suffices to correct it by the cosine in Eq. (14). Notice that Eq. (15) includes the possible additional branching-point mode in Eq. (13) (when not suppressed by *k*), obtained simply by setting *λ* = 0. It is convenient to compute its limiting values:

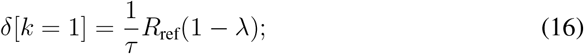

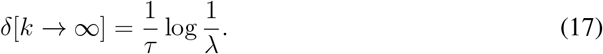

The first implication is that if the generation time has *k ≈* 1 (i.e., VAR[*τ*] *≈ τ* ^2^) then the subdominant eigenmodes will decay faster in more transmissible epidemics: the larger is *R*_ref_, the faster *R*_estim_ converges to *R*_ref_. Instead, when the dispersion of the generation time decreases (*k* increases) the decay rate of the modes no longer depends on the overall transmissibility of the disease (Eq. (17)). Also, by imposing the condition *δ*[*k → ∞*] *< δ*[*k* = 1] one finds a cutoff value

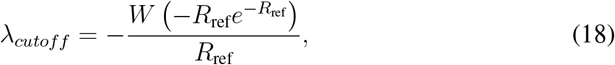

where *W* is the principal branch of the Lambert’s *W* function: increasing *k* will decrease the *δ* of the modes for which *λ > λ*_*cutoff*_ (and thus increase the contribution of each eigenmode). It will instead decrease the contribution of those for which *λ < λ*_*cutoff*_. Below the epidemic threshold (*R*_ref_ *<* 1) one has *λ*_*cutoff*_ = 1, meaning that underdispersed generation times (large *k*) will lead to faster convergence of *R*_estim_ to *R*_ref_ because all the modes are increasingly suppressed. Conversely, above the epidemic threshold, under-dispersion will amplify the effect of many eigenmodes, as *λ*_*cutoff*_ is fairly small even for moderately big *R*_ref_: e.g., *λ*_*cutoff*_ = 0.2 for *R*_ref_ = 2. The behavior at small *k* (strong overdispersion) has an additional feature. *dδ/dk* is always negative close to *k* = 0 if *R*_ref_ *>* 1 so that very strong values of overdispersion will always decrease eigenmode contribution even if *λ < λ*_*cutoff*_. Finally, the dispersion of the generation time can increase the number of relevant exponential terms for each eigenmode by interacting with the spatial network. Equation (14) shows that, when *k* is large, terms other than *m* = 0 can provide nonnegigible contributions, as cos(2*π/k*) increases as *k* increases. Underdispersed generation thus further amplify the contribution of subdominant eigenmodes and slow the convergence of *R*_estim_ to *R*_ref_, also increasing the number of relevant exponential terms, with increasing likelihood of seeing complex oscillatory behaviors of *R*_estim_ around *R*_ref_.

To confirm and illustrate the effect of the distribution of the generation time on the dynamical convergence of the epidemic to its dominant eigenmode, we simulated the spread of a respiratory pathogen in Spain. We used Colocation Maps by Meta [51] to parametrize the operator **R** [19, 27], identifying Spanish provinces as spatial communities. More details are available in Appendix D. Figure 1(B,D) compare the evolution in time of the value of the reproduction ratio estimated from surveillance (*R*_ref_) and the reference value (*R*_ref_) for two hypothetical epidemics started in Spain’s capital Madrid. They were parametrized using COVID-19 (original strain) and pandemic H1N1 except for the dispersion parameter *k* which was explored, to show the previously discussed slowdown of the convergence to *R*_ref_ due to an underdispersed generation time.

Figure 2 further explores the interaction between spatial dynamics and generation time. Specifically, Fig. 2A confirms the existence of the structural cutoff (Eq. (18)) by showing how the effect of *k* on the time of convergence to *R*_ref_ changes with the overall transmissibility, in the case of a COVID-19-like epidemic starting in Madrid. As *R*_ref_ is close to the epidemic threshold, convergence is faster for higher *k* values. But as *R*_ref_ is slightly above the critical value convergence slows down dramatically as *k* increases, as one or more subdominant modes in the Spanish network find themselves above *λ*_*cutoff*_. The figure also confirms that at large *k* the time of convergence depends weakly on overall transmissibility (see Eq. (17)). Figure 2(B-D) explore the effect on time of convergence of a generic subdominant mode when it varies in magnitude. As *k* = 1 (exponential distribution) Fig. 2C shows the linear relationship between convergence and *R*_ref_, *λ* found in Eq. (16). Fig. 2B instead confirms that a highly overdispersed generation time (*k* = 0.1 in Fig. 2B) sharply flattens the contribution of all eigenmodes below *λ*_*cutoff*_ to zero dramatically slowing down convergence. But as *R*_ref_ increases, those modes become larger than *λ*_*cutoff*_ and at that point their weight is sharply increased, abruptly decreasing convergence time. Here, convergence time is sensitive almost exclusively to *R*_ref_, which determines – through *λ*_*cutoff*_ – whether the contribution of the subdominant modes is pushed towards zero or toward one. The situation is inverted in Fig. 2D. When the generation time is underdispersed (large *k*) we again see the effect of Eq. (17): the contribution of the sub-dominant modes does not depend on *R*_ref_ and the time of convergence is weakly sensitive to the overall transmissibility.

**Fig.2.**
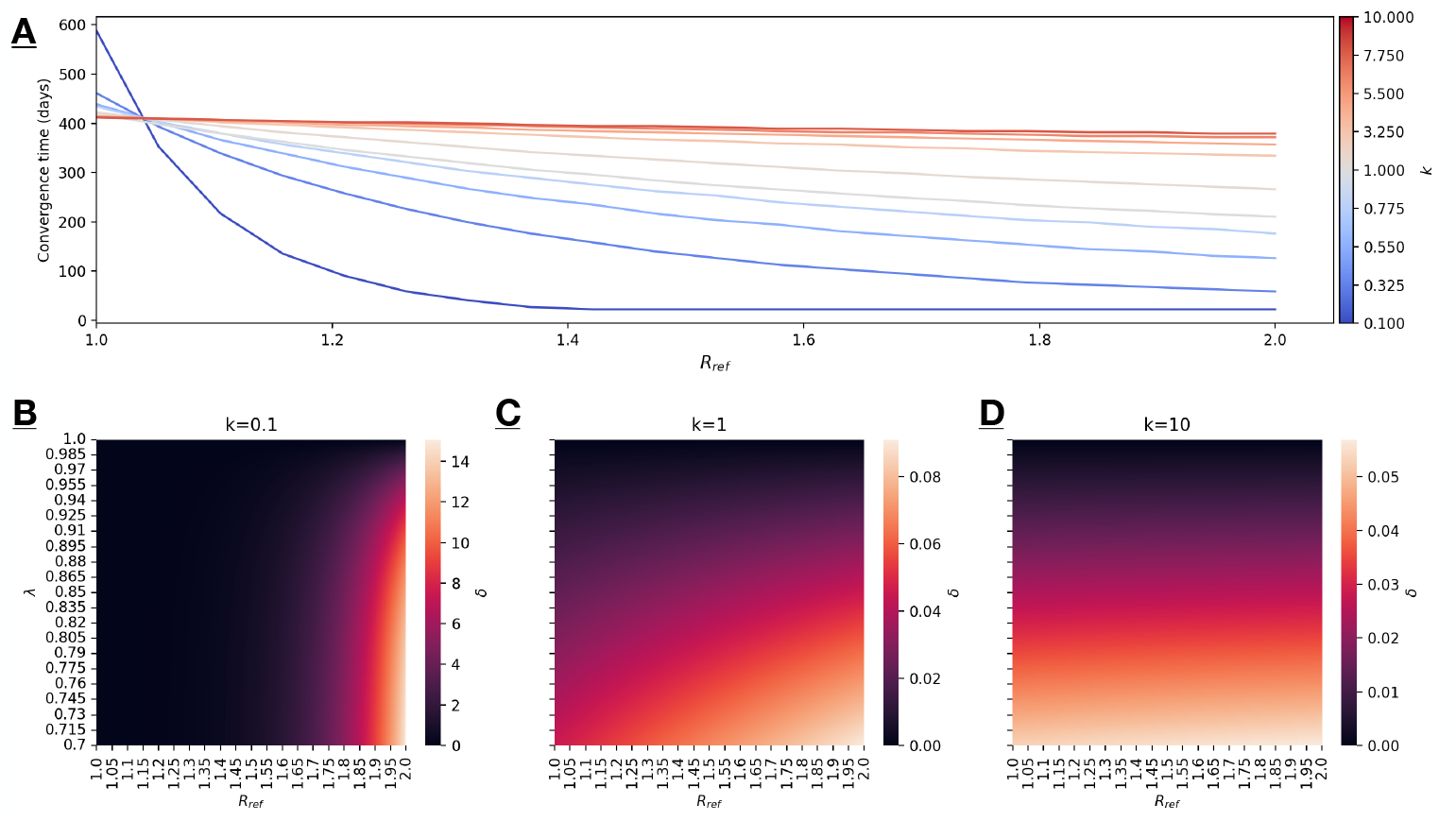
In panel **A**, the convergence time of *R*_estim_ towards *R*_ref_ is shown as a function of *R*_ref_. The generation time distribution is modeled as a gamma distribution with the mean set to the estimate for COVID-19 (original strain) [49] and varying shape parameter *k*. Panels **B, C**, and **D** show heatmaps of the *δ* rate as a function of *R*_ref_ and the *λ* parameter for *k* = 0.1, *k* = 1, and *k* = 10, respectively.

### 4.2 When the generation time is not correctly specified

We model this case by assuming that *ζ*_*estim*_ is a gamma distribution with mean *τ*_*estim*_ and *k*_*estim*_ which may be different from the “true” values *τ, k*. We will focus on *R*_corr_, which we proved to be an unbiased estimator of *R*_ref_ if *τ, k* are correctly specified.

From Eq. (9) we can compute the scalar Green’s function for 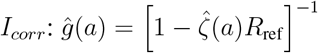, which is simply the component of Eq. (5) proportional to **v**. Assuming, for simplicity, no source term beyond *t* = 0 (*J*_*corr*_(*t*) = *δ*(*t*)*j*_0_), then

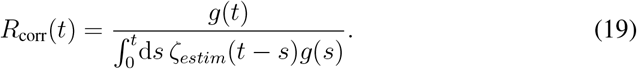

The singularities of *ĝ*(*a*) are those in Eq. (11), with each residue

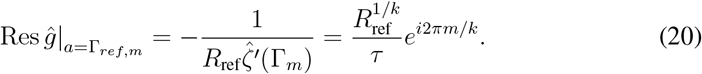

As in the case of the full Green’s function, we write

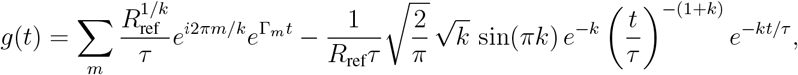

where the second term accounts for the branch point discussed in Appendix B. Since this contribution, when present, always decays in time, we will neglect it in what follows. To estimate the denominator in Eq. (19) we first consider its Laplace transform: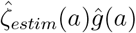. We can neglect the singularities coming from the first factor 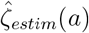 as they decay fast: their real part must be negative given that *ζ*_*estim*_(*t*) *→* 0 as *t → ∞*, being a probability density function. Therefore the role of 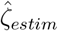 is to correct the residues of the already-considered singularities, giving the final formula:

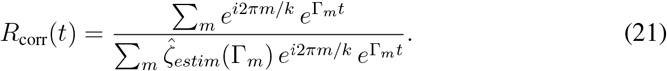

Note that when 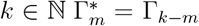 and Γ_*k/*2_ *∈* ℝ when *k* is even, so that this expression is real. If *k∉* ℕ we know it to be an approximation so that residual imaginary terms may linger. As a safety check, one can set *ζ*_*estim*_ = *ζ* in Eq. (21). Then 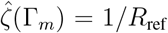 by definition, so that *R*_corr_(*t*) = *R*_ref_ correctly at all time. When this is not the case, the combination of modes will bias the estimate of *R*_ref_ even when the incidence has been corrected by **v**^*\**^, with a possibly oscillating sign of the error (underestimation and overestimation of *R*_ref_). But most interesting is the asymptotic value of Equation (21), when *t → ∞*. There, only *m* = 0 counts, so that

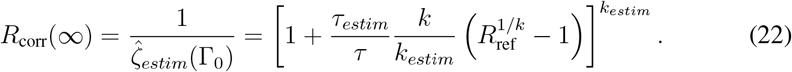

This value is generally different from *R*_ref_, meaning that the bias coming from misestimating the generation time distribution does not decay to zero in time, unlike the one coming from neglecting or misestimating the spatial structure. This happens because unlike the structural bias which is self-correcting as the dynamics tends to the spatial equilibrium, the bias from the generation time is not. Its origin and magnitude, however, do vary according to dynamical parameters. Expanding Eq. (22) for *R*_ref_ *≈* 1, i.e., when the epidemic is close to the epidemic threshold, gives the following

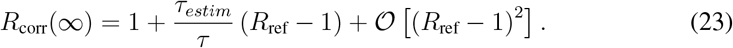

This provides two important pieces of information: first, when the epidemic is close-to-critical, the error coming from misestimating the generation time is small. Second, accurately knowing the average generation time is enough as Eq. (23) does not depend on *k*: This is notable because estimating the average generation time is easier than estimating its dispersion [52, 53]. However, when *R*_ref_ is far from the critical point, Fig. 3 shows that knowing *τ* is not enough, and an accurate estimate of *k* is needed to ensure that *R*_corr_ measures *R*_ref_.

**Fig.3.**
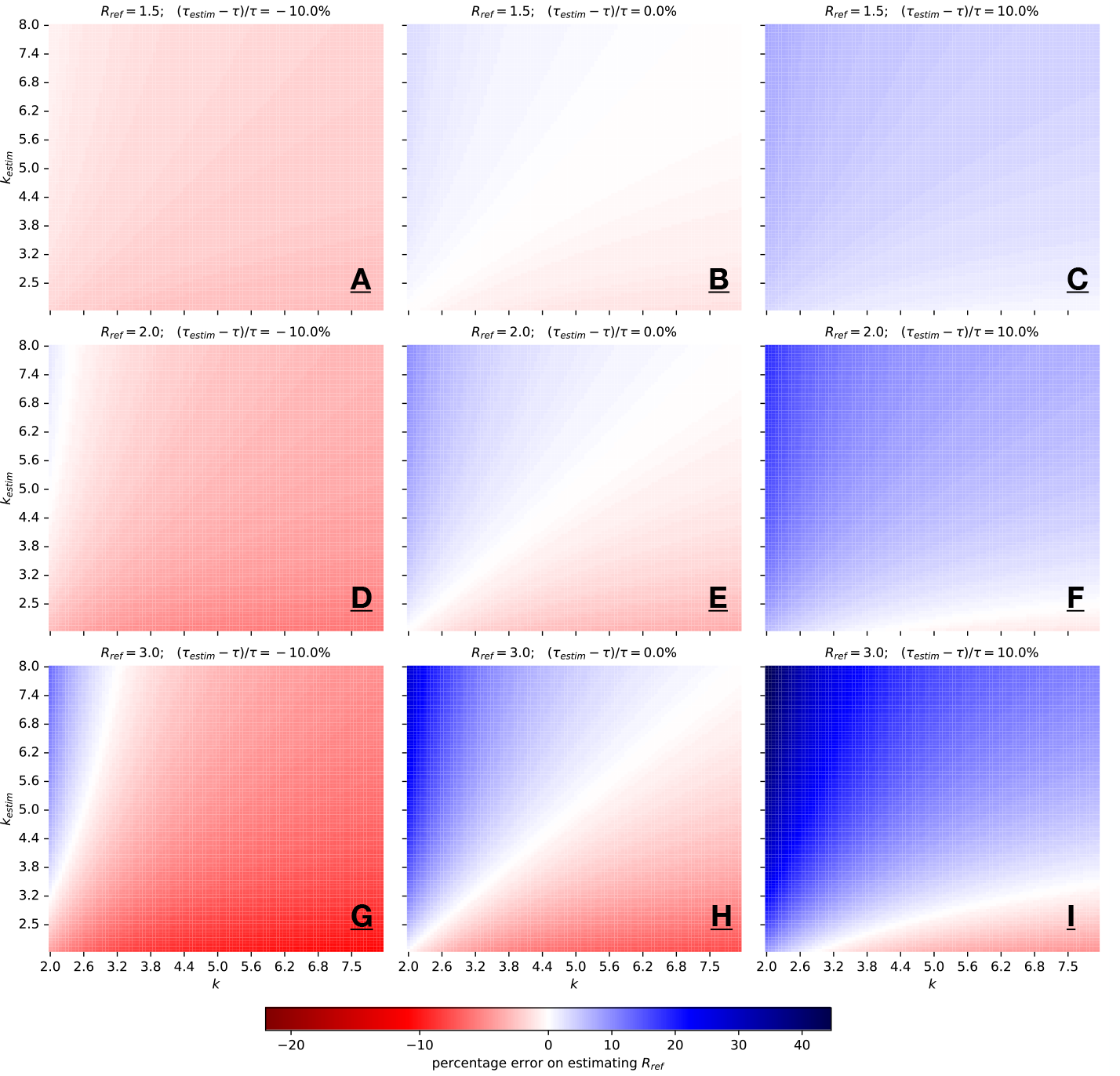
Effect of inaccurately measuring the generation time distribution on the estimate of the reproduction ratio. Each panel shows the relative percentage error on the estimates of the reproduction ratio. Rows indicate the actual shape parameter *k* of the generation time distribution, columns the one used in the estimate. Panels have different values of the reference reproduction ratio *R*_ref_ and of the relative difference between the actual mean generation time and the one used to computer the reproduction ratio. The color bar shows the relative difference between the estimated reproduction ratio and *R*_ref_ asymptotically at large time.

Figure 4 shows the error made by estimating the reproduction ratio of one historical epidemic of respiratory pathogen, using the generation time parametrized from another epidemic. The magnitudes are substantial, and this is a problem because this is often done in the absence of reliable data [54, 55].

**Fig.4.**
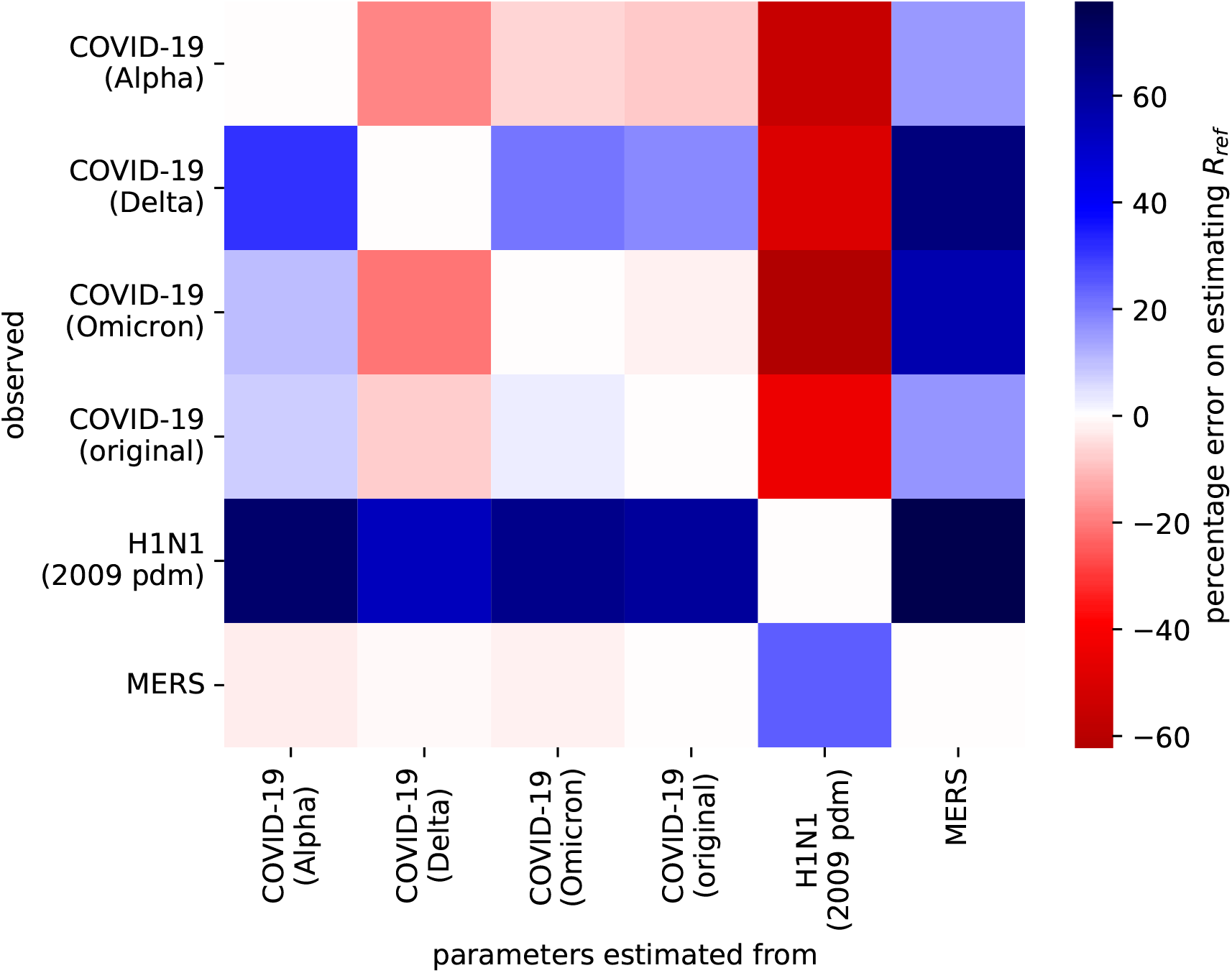
Impact of sharing estimates of the generation time across epidemics. The figure shows the relative percentage error on the estimates of the reproduction ratio which is made when estimating *R* for the epidemic indicated in the column using parameters from the epidemic indicated in the row. The parameters used are *R*_ref_ = 3.0, *τ* = 6.60, *k* = 2.49 for the original strain of COVID-19 [49, 56], *R*_ref_ = 4.5, *τ* = 7.12, *k* = 2.53 for its Alpha variant [57, 58], *R*_ref_ = 5.0, *τ* = 6.52, *k* = 1.87 for its Delta variant [57, 59], *R*_ref_ = 8.2, *τ* = 6.84, *k* = 2.39 for its Omicron variant [60], and *R*_ref_ = 0.7, *τ* = 6.80, *k* = 4.85 for 2012–2013 Middle East Respiratory Syndrome (MERS) [61]. The color bar shows the relative difference between the estimated reproduction ratio and *R*_ref_ asymptotically at large time.

## 5 Perturbations

We will now use the formalism developed so far to study the effect of a short-term change in the reproduction operator on the spatial dynamics, and on the estimate of the reproduction ratio. We assume that the system is governed for a long time by an unperturbed **R**, so that the dynamics has evolved to the spatial equilibrium **v**. Practically, this means that the system is prepared in the state **v** at *t* = 0. In the absence of perturbations, the spatial distribution of incident infections will remain **v** indefinitely. We wish to measure how much a perturbation changes the spatial distribution, and how long it takes for the system to revert back to **v** after the perturbation. This information is relevant because the distribution of infections describes the dynamics of the epidemic in space, and it impacts the ability of public health to correctly estimate epidemic evolution, as we have seen. Specifically, we will assume that the reproduction operator changes to **R**_*p*_ in the interval *t ∈* [0, *t*_*p*_], and then reverts back to **R**. This simplified system can model the effect of changes in the spatial mixing of communities due to exhogenous perturbations: extreme weather [26–29], public health interventions such as mobility restrictions [31], seasonal patterns such as holidays [33, 34].

We identify the spectral structure of **R**_*p*_ with the subscript _*p*_, so that, for instance, its reference reproduction ratio and spatial equilibrium (Perron eigenvector) are *R*_ref,*p*_, **v**_*p*_. Analogously, we identify the Green’s function of the perturbed evolution as 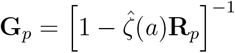. We assume that the perturbation affects the overall transmissibility and spatial epidemic structure but does not change the generation time distribution.

Appendix E proves that the Laplace transform of **I**(*t*) after the perturbation is

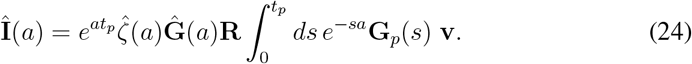

The first exponential simply shifts the exponential growth to start at *t*_*p*_. The term **Ĝ** (*a*) is the only one contributing with singularities, correctly expressing the fact that the asymptotic behavior (large *t*) will be determined by the eigenmodes of the unperturbed propagation [43]: each of them will decay at a relative rate given by Eq. (14). The last term in Eq. (24) encodes the effect of the perturbation as it computes the weight of each sub-dominant eigenmode attributable to the perturbation. To isolate each weight we multiply from the left by 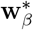 and, as before, compute the residue. Assuming again for simplicity real positive eigenvalues, for the mode Γ_*β,m*_ the residue is

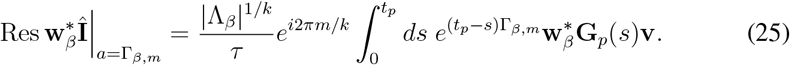

The term 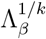 is the familiar weight (Eq. 13) while the last term is the contribution of the perturbation. It shows that the most disruptive perturbations are those whose propagators transform **v** into **w**_*β*_, where *β* is the one with the smallest decay rate *δ*. Note, however, that this transformation will never be exact as **w**_*β*_ is not a Perron vector and thus not a suitable spatial distribution, but will maximize the scalar product with 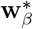. The simplest way to achieve it is that **R**_*p*_ contains a term approximately proportional to **w**_*β*_**v**^*\**^. Let us consider this term in the context of weakly coupled systems, whereby transmission occurs predominantly within communities or within clusters of communities, translating into diagonally dominant or block-diagonally dominants reproduction operators (*R*_*ii*_ *>*∑ _*j*≠*i*_ *R*_*ji*_). There, each eigenvector is mostly concentrated into one or few communities, and the outer product 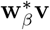 indicates, in the context of a perturbation, an anomalous increase (or decrease) in the transmission potential between the communities where **w**_*β*_, **v** are concentrated. This was observed, for example, in the aftermath of natural disasters where aid-driven mobility increases contacts among residents of communities which can be far apart [27], or in the time leading to a major movement restriction such as the first COVID-19-driven lockdown in France, when people spontaneously and massively relocated outside major urban centers during the time between policy announcement and policy enforcement [31]. More routinely, it can also happen during holiday periods when people spend time in crowded destinations away from their normal residence [33, 34]. These examples use administrative units as communities; similar effect may however occur across spatial scales, when perturbations change contacts between the distinct *mobility containers* that are known to exist at multiple scales [62].

As a final observation, we wish to determine the impact of the generation time distribution on the effect of the perturbation. To do this, we assume the perturbation to last for a relative short time and expand the perturbation term in *t*_*p*_ (see Appendix F). We find that for up to *k ≈* 1

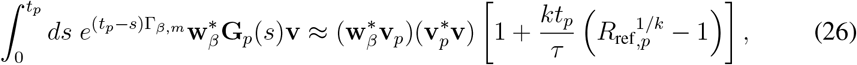

where, for clarity, we have kept only the leading mode of the perturbed propagation. For exponentially-distributed generation times (*k* = 1) the effect of the perturbation grows linearly with the transmissibility during the perturbation, while overdispersed generation times (*k <* 1) enhance this through a sublinear relation with small argument.

For underdispersed generation times (*k ≫* 1):

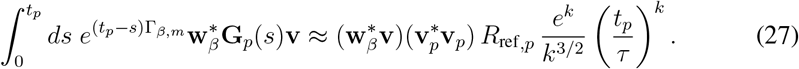

The exponential dependency on *k* as well as the leading order 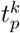 show that underdispersion substantially enhances the effect of the perturbation.

To sum up, this means that the interaction between generation time and spatial structure again affects epidemic dynamics by enhancing the effect of changes in spatial transmission patterns when *τ* is both underdispersed and overdispersed with respect to the exponential distribution. The fact that most epidemics of respiratory pathogens feature underdispersed generation times (see Fig. 4) and, more generally, generation times compatible with *k* = 1 are rarely observed, implies that the effect of perturbations, even if relatively short (weeks) [27, 31, 63], may substantially change the spatial structure of the epidemic and compromise our ability to correctly evaluate epidemic trends using surveillance data.

## 6 Conclusion

In this study, we developed a theoretical framework linking the effect of the distribution of the generation time and the spatial contact structure on epidemic dynamics, and the implications for epidemiological surveillance.

We showed that the generation time distribution interacts nontrivially with the spatial coupling network to determine epidemic evolution and, in particular, the rate and manner in which the system approaches the equilibrium spatial distribution of infections. Our findings show that the dispersion of the generation time around its mean modulates the contribution of subdominant epidemic modes, thereby altering the time required for surveillance-based estimates of *R* to converge to their reference value. For epidemics close to criticality, overdispersed generation times slow convergence, while for transmission well above the epidemic threshold, underdispersed generation times —-typical of many respiratory pathogens -— can markedly delay convergence, leading to long-lived bias in surveillance-derived *R*.

Furthermore, we found that the previously introduced correction to incidence data, which removes the bias due to the spatial structure, remains valid for arbitrary generation time distributions only if both the mean and the shape of the distribution are accurately known. When *R* is close to *R* = 1, errors from misspecifying the generation time are small and knowledge of the mean *τ* suffices. But for *R* well above the critical point, mis-estimation of either the mean or the dispersion of the generation time induces systematic errors that are not removed by the spatial correction and do not decay in time.

Finally, we applied the framework to short perturbations in epidemic dynamics, such as those arising from exogenous factors (e.g., extreme weather events, mass gatherings, sudden mobility restrictions) as well as endogenous changes (e.g., behavioural changes). We found that realistic dispersion values can substantially amplify the impact of such perturbations and increase the time needed by the epidemic to return to the unperturbed equilibrium. As a result, even in regimes where surveillance-based estimates *R* would otherwise be reliable, abrupt short-term changes can induce biases in *R* estimates that persist well beyond the duration of the perturbation.

We illustrated these effects in a case study of a respiratory pathogen in Spain, where colocation-based mobility data allowed us to quantify the interaction between generation time distribution and spatial structure, and their effects on the estimate of *R*, under realistic contact patterns.

In conclusion, these findings strengthen the theoretical basis, and provide operational criteria for public health practice, to interpret surveillance-derived estimates of epidemic evolution, and to delineate the epidemiological and statistical conditions under which such estimates remain robust, and the availability of corrections.

## Data Availability

The colocation maps used are available at https://dataforgood.facebook.com/dfg/ tools/colocation-maps

## A Asymptotic evolution

Equation (5) implies that, as *t → ∞*,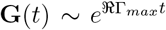, where Γ_*max*_ is the singularity of **Ĝ** (*a*) with the largest real part. The singularites solve the following equations:

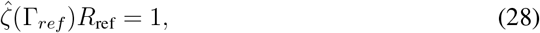

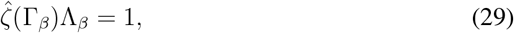

whose explicit formula in the case of a gamma-distributed generation time appears in Eq. 11 and Eq. 12. Here, however, we consider a completely general generation time distribution. This is a quite cumbersome proof of something apparently trivial, i.e., that the dominant eigenmode of **R** is indeed what determines the asymptotic epidemic behavior. We report it nonetheless, to prove that changes in the generation time distribution can change how the system approaches the asymptotic equilibrium spatial distribution **v** but cannot change it, as it is exclusively determined by the spatial network structure.

We prove here that Γ_*max*_ is the real solution of Eq. 28. To do this, we study 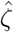. Clearly, 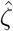 is a real function for *a ∈* ℝ. The requirement that *ζ*(*t*) *≥* 0 for *t ∈* ℝ_+_ implies that 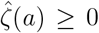 for *a ∈* ℝ _+_. 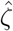 is also monotonically decreasing on ℝ_+_ because 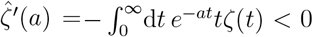 for *a ∈* ℝ _+_. Its (possibly multivalued) inverse 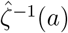 is thus also monotonically decreasing:

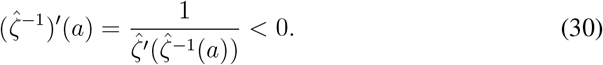

Now we need to prove that *ℜ*(Γ_*β*_) *<* Γ_*ref*_ *∀β*.

**Case 1:** Λ_*β*_ *∈* ℝ

Since **R** is nonnegative irreducible matrix then Λ_*β*_ *< R*_ref_ by Perron-Frobenius theorem.

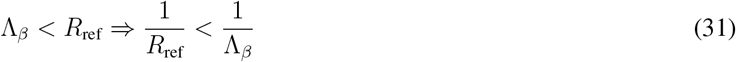

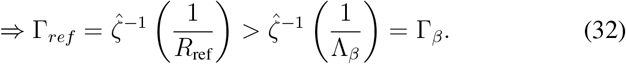

This proves that Γ_*ref*_ *>* Γ_*beta*_. We dropped *ℜ* because being *ζ*^^*−*1^(*a*) is real if the argument is real.

**Case 2:** Λ_*β*_ *∈*ℂ *\* ℝ

Given that 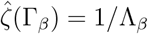, then

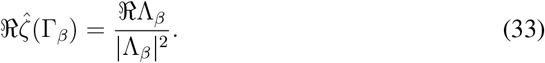

We now prove the following inequality:

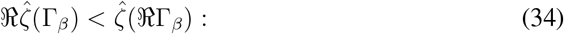

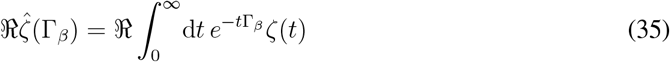

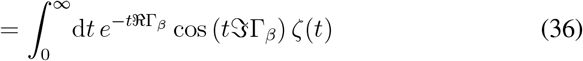

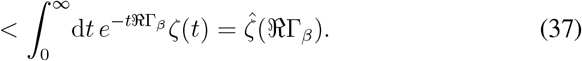

Combining Eq. 33 and Eq. 34:

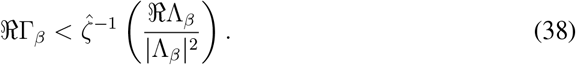

Then to prove that Γ _*ref*_ *> ℜ*Γ_*β*_ we just need to prove that 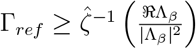:

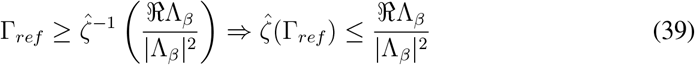

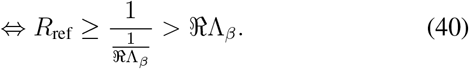

## B Gamma-distributed generation time

We assume the generation time *t* to be distributed as a gamma distribution, defined on [0, *∞*) and with the following probability density function:

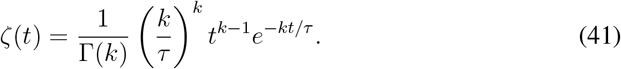

*τ* is the mean generation time and *k* is the shape parameter tuning dispersion:

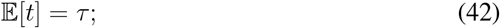

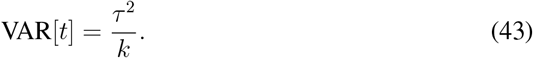

This distribution has the following Laplace transform:

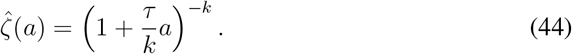

### B.1 Green’s function

We study the time evolution of the Green’s function **G**(*t*) from its Laplace transform in the case of gamma-distributed generation time. We identify the generic term making up Eq. (5) as follows:

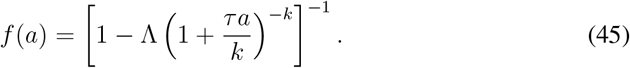

For simplicity we define a shifted variable: *x* = 1 + *τa/k* and a function 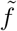 such that 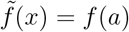:

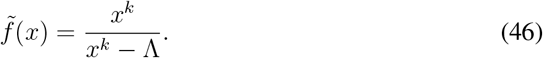

In general, *k ∈* ℝ. Here, however, we will assume that *k ∈*ℚ. This cause no loss of generality as estimates of the dispersion of the generation time have always nonzero uncertainty, so they can always be approximated arbitrarily well with a rational number given that ℚ is dense in ℝ. Candidate singularities of 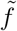 are the solutions of *x*^*k*^ *−* Λ = 0 and *x* = 0, the latter existing only if *k∉* N and being a branch point. We will first prove that the former singularities are a finite number of simple poles and compute their residue.

Let *k* = *p/q* with *p, q ∈*ℕ coprime. Then the solutions of *x*^*k*^ *−* Λ = 0 are a subset of the solutions of *x*^*p*^ *−* Λ^*q*^ = 0. These latter are

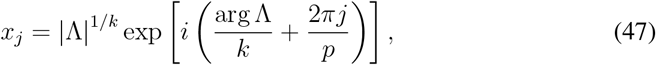

with *j* = 0, 1, *· · ·, p −* 1. Among these, *x*_*j*_ is a solution of *x*^*k*^ *−* Λ = 0 only if *q* divides *j*. This means that the solutions of *x*^*k*^ *−* Λ = 0 can be enumerated as follows:

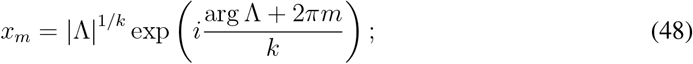

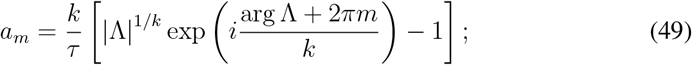

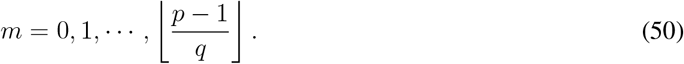

Here, we also displayed the singularities shifted back to *a*. We identify the numerator and the denominator in Eq. (46):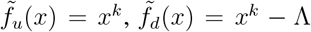. Now 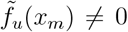 and 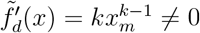 so 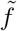 has simple poles in *x*_*m*_. Also,

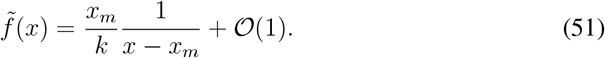

Going back to *a*, this means that *f* has simple poles in *a*_*m*_ and

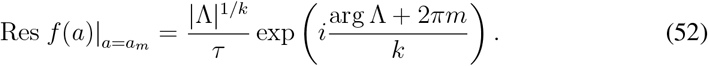

Each of these simple poles contributes to an exponential growth in each term of **G**(*t*) with rate Γ_*m*_ = *a*_*m*_ and weight equal to Res 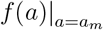.

We now turn to the branch point in *x* = 0 or, equivalently, in *a* = *−k/τ*. Close to the branch point, 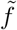 behaves as

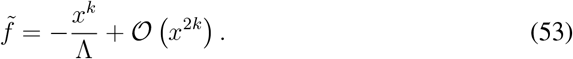

Performing the inverse Laplace transform of this local expansion and shifting back to *a* one finds that the branch point contributes to the time evolution of each term of **G**(*t*) with the following:

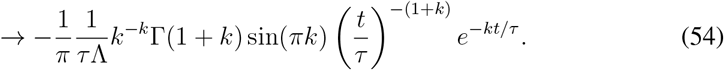

Given that the branch point exists only if *k∉*ℕ, the term sin(*πk*) correctly suppresses it when *k ∈*ℕ. For *k ≫* 1 we can replace Euler’s Gamma with its Stirling approximation (see Fig. 5):

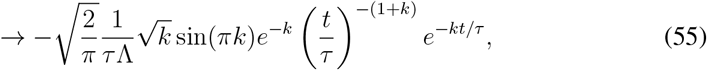

which means that this contribution is exponentially suppressed at large *k*, as proven also by Fig. 5. This figure also shows that this contribution can be relevant basically for *k* around 1*/*2 and possibly 3*/*2, when the *k*-dependent coefficient peaks and the exponential decay in time *e*^*−kt/τ*^, whose rate depends linearly on *k*, is still small.

**Fig.5.**
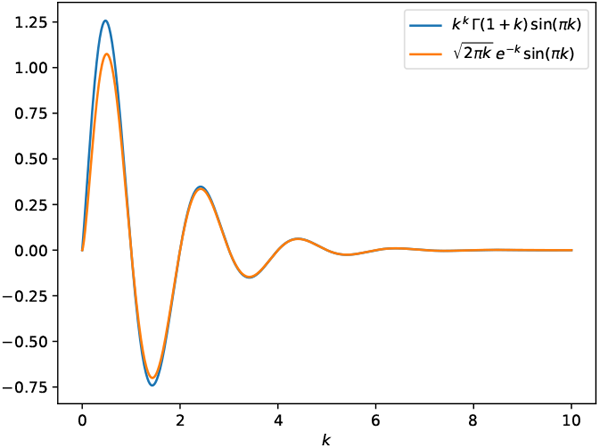
Evaluation of the *k*-dependent term of the contribution from the branch point, both it its exact form (Eq. (54)) and with Stirling’s approximation (Eq. (55)).

## C An identity with G

Proof that

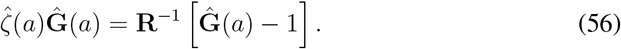

Note that here **R**^*−*1^ can be formal if **R**^*−*1^ is not invertible as the inverse disappears at the end of the calculations.

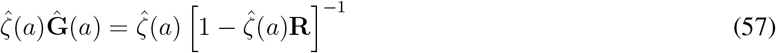

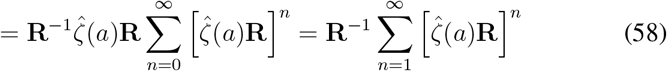

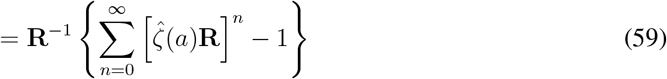

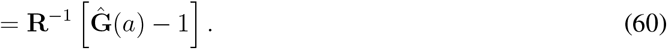

## D Numerical simulations

### D.1 Parametrization of the reproduction operator

To reconstruct the spatially structured reproduction operator, we use Colocation Maps from Meta, which quantify the probability *p*_*ij*_ that a randomly selected resident of community *i* and one of community *j* are found colocated during a 5-minute interval within a given week. These maps allow us to stratify by type of contact (within household or community). For our purposes, we only focused on community contacts. These maps are provided at the administrative level of Spanish provinces in April 2023. Similarly to what we did in Ref. [19, 27], we define the unnormalized contact rate matrix as *M*_*ij*_ = *p*_*ij*_*N*_*j*_, where *N*_*i*_ is the resident population of community *i*, obtained from census data [64]. Finally, we parametrize the entries of the reproduction operator as

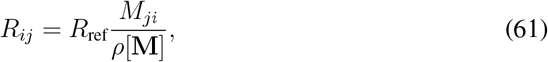

where *ρ* is the spectral radius, so that the scalar coefficient tuning **R** is exactly *R*_ref_. This operator captures both within-community (*i* = *j*) and between-community (*i≠ j*) transmission potential.

### D.2 Numerical solutions

We solved numerically the Volterra equations by discretizing it in a certain number of steps. We fixed a timestep *dt* = 0.04 *d*, an initial condition **v**_0_ where we considered the first case reported to be introduced in the spatial community of Madrid and an observation interval *T*. We then solved the equation for every timestep and for every community thus obtaining a vector of cases of dimensions *N × T/dt* where *N* is the number of spatial communities and *T/dt* is the number of timesteps considered. To obtain *R*_estim_, we computed the total incidence per timestep as the sum across all communities at a specific one, *I*_tot_(*t*) = Σ_*i*_ *I*_*i*_(*t*). From this, we exploited the relation:

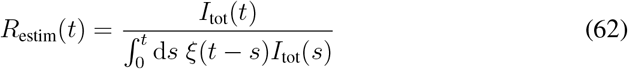

To extract the time of convergence, we firstly computed the relative variation between *R*_estim_ and *R*_ref_, called it Δ(*t*). Then, we defined the time of convergence as the smallest *t*^*\**^ for which Δ(*t*^*\**^) *< ϵ* where *ϵ* = 0.01 arbitrarily. To ensure this value would be representative for the long-time convergence of the estimate, we disregarded any result before *t* = 20 *d*. Considering a gamma distribution as the generation time distribution, we explored these 2 values at varying shape and for different values of the mean.

## E Evolution after spatial perturbation

The evolution of the epidemic with the perturbation is described by the following:

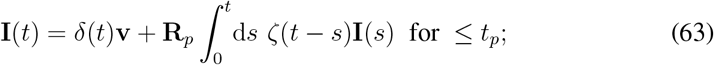

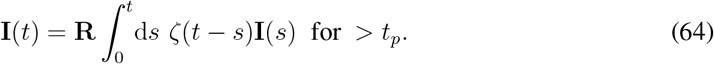

Equation (63) has the following solution: **I**(*t*) = **G**_*p*_(*t*)**v**. Equation (64) can be decomposed as follows:

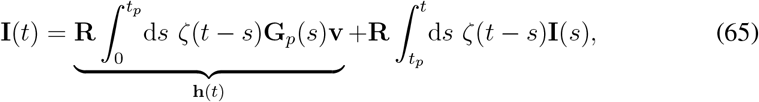

where we replaced **I**(*s*) in the first integral with the solution at *t ≤ t*_*p*_. Now this first integral (**h**(*t*)) no longer contains the unknown and acts as a source term. We define the shifted variables 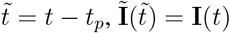 and 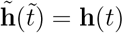 and rewrite:

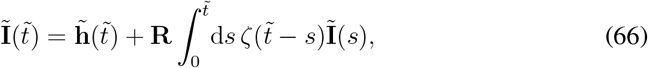

whose solution for 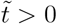 is

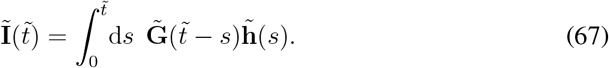

We perform the Laplace transform of this last equation noting that the time shifting generates 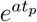 terms:

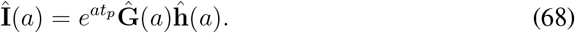

Performing that the Laplace transform of **h**, one gets Eq. (24).

## F Short-time expansion of the perturbation

To compute the short-term expansion in the time domain we compute the asymptotic (large *a*) expansion in the Laplace domain:

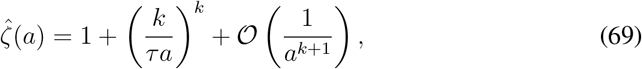

which leads to, for each term in the Green’s function (see Eq. (5)):

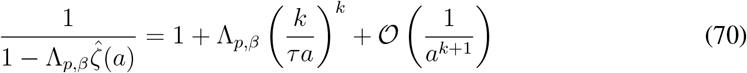

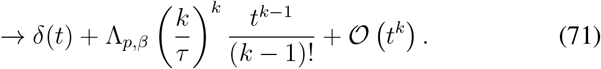

This last term appears in the integral:

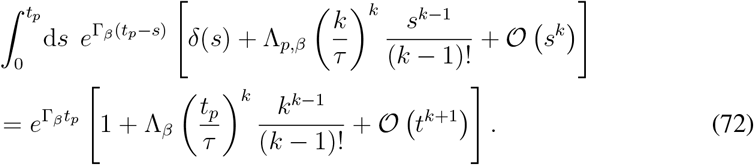

We note that, for *k* large enough,

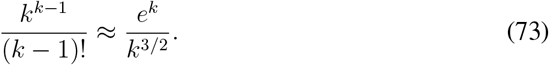

